# Genetically influenced tobacco and alcohol use behaviors impact erythroid trait variation

**DOI:** 10.1101/2023.05.01.23289329

**Authors:** Shriya Shivakumar, Madison B Wilken, Victor Tsao, Bárbara D. Bitarello, Christopher S Thom

**Author notes:** Correspondence: Christopher S Thom, MD, PhD, Children’s Hospital of Philadelphia, Colket Translational Research Bldg, Room 10-052, 3501 Civic Center Blvd, Philadelphia, PA 19104.

## Abstract

Genome wide association studies (GWAS) have associated thousands of loci with quantitative human blood trait variation. Blood trait associated loci and related genes may regulate blood cell-intrinsic biological processes, or alternatively impact blood cell development and function via systemic factors and disease processes. Clinical observations linking behaviors like tobacco or alcohol use with altered blood traits can be subject to bias, and these trait relationships have not been systematically explored at the genetic level. Using a Mendelian randomization (MR) framework, we confirmed causal effects of smoking and drinking that were largely confined to the erythroid lineage. Using multivariable MR and causal mediation analyses, we confirmed that an increased genetic predisposition to smoke tobacco was associated with increased alcohol intake, indirectly decreasing red blood cell count and related erythroid traits. These findings demonstrate a novel role for genetically influenced behaviors in determining human blood traits, revealing opportunities to dissect related pathways and mechanisms that influence hematopoiesis.

## Introduction

Tobacco and alcohol use are prevalent behaviors that have many detrimental effects on human health, with >85% of U.S. adults over the age of 18 reporting alcohol use and >12% (>30 million) U.S. adults actively smoking cigarettes (U.S. CDC, 2022; U.S. NIAAA, 2022). While associated cardiovascular disease risks are well documented, effects on many organ systems are not well understood. For example, clinical observational studies have linked tobacco and/or alcohol use with altered blood cell traits (Nordenberg et al., 1990; Sunyer et al., 1996; Ballard, 1997; Asgary et al., 2005; Malenica et al., 2017). These findings may reflect effects on blood cell development or function, with potential broad implications for human health. However, prior studies have reported conflicting blood trait effects. A complex interplay between physiologic mechanisms may link tobacco use with anemia (Leifert, 2008), although multiple studies have associated smoking with higher hemoglobin levels (Nordenberg et al., 1990; Malenica et al., 2017). Alcohol use has generally been associated with anemia, via effects on erythrocyte production, metabolism, and function (Ballard, 1997). However, these conclusions are largely based on observational studies that are subject to bias from unmeasured or unrecognized confounding factors. We hypothesized that genetic methods might be used to clarify the directional effects of smoking and alcohol use behaviors on blood traits.

Prior blood trait genome wide association studies (GWAS) adjusted for tobacco and alcohol use behaviors given effects on multiple blood traits (Astle et al., 2016). The identities of affected traits, as well as the directional effects of these behaviors on specific blood traits, are unknown. More recent GWAS were not adjusted for tobacco or alcohol use (Chen et al., 2020; Vuckovic et al., 2020), providing an opportunity to analyze genetically determined effects of these behaviors on blood traits. We reasoned that such studies would have increased power to discover associations between behavioral effects on genetically determined blood traits, compared with a prior study (Pedersen et al., 2019).

A key outstanding question is if and how behavioral factors influence genetically determined blood trait variation. Such influences and associations are potentially relevant for understanding genetic determinants of blood development and disease phenotypes. Here, we used a Mendelian randomization (MR) framework to establish causal effects of tobacco and alcohol use on blood traits. MR leverages random single nucleotide polymorphism (SNP) allocation at meiosis to make causal inferences about the effects of a genetically determined exposure trait on an outcome phenotype (Burgess et al., 2020). Multivariable MR (MVMR) can extend this method to identify mediating or confounding traits to explain causal effects (Sanderson et al., 2019). In some cases, we identified behavioral effects ran counter to clinical observations and widely held clinical assumptions. Using MVMR and formal mediation strategies, we identified relevant trait relationships to explain unexpected erythroid trait relationships and determine correct causal directions. Our results clarify genetic effects of tobacco and alcohol use behaviors on quantitative blood traits.

## Results

### Increased genetically determined odds of smoking initiation decrease hemoglobin, hematocrit, and red blood cell count

We conducted two sample Mendelian randomization (MR) experiments to ascertain if genetically determined smoking initiation (SmkInit) risk impacted one or more of 15 quantitative blood traits (**Fig. 1A, Supplementary Fig. 1**, and **Supplementary Table 1**). An instrumental variable comprising 113 linkage-independent SNPs significantly associated with SmkInit negatively impacted erythroid traits, including quantitative measures of hemoglobin (HGB), hematocrit (HCT), and red blood cell counts (RBC) by the inverse variance weighted (IVW) method (p<0.05 for each blood trait). These experiments revealed non-significant MR Egger intercept values, arguing against horizontal pleiotropy (all p>0.05). Sensitivity analyses using weighted median and MR Egger methods showed directionally consistent results (**Supplementary Fig. 2**), and reverse causality experiments also supported a directional effect from smoking on red blood cell (erythroid) traits (**Supplementary Fig. 3**). The effects of SmkInit did not extend to quantitative platelet and white blood cell characteristics (**Fig. 1A**). The instrumental variables used for these experiments, and all other experiments in this manuscript, were not subject to weak instrument bias as defined by F statistics > 10 (**Supplementary Table 2**) (Burgess and Thompson, 2011).

**Figure 1.**
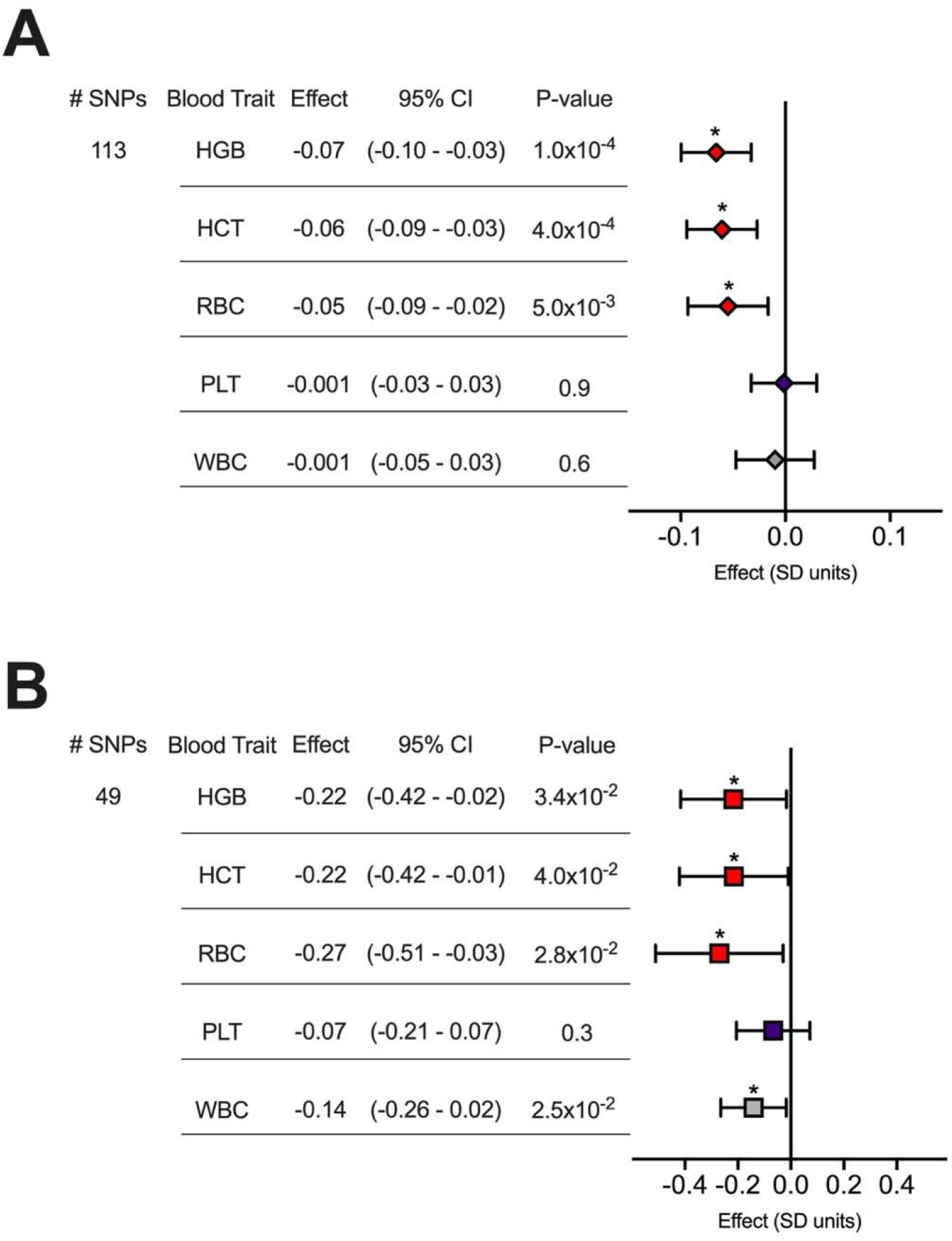
Smoking initiation (SmkInit) and alcohol use (Drinks) negatively impact genetically determined hemoglobin (HGB), hematocrit (HCT), and red blood cell count (RBC). (**A-B**) MR results using the inverse variance weighted method are shown, with values for Effect sizes on the indicated blood traits reflecting (**A**) a 2-fold increase in SmkInit risk or (**B**) a 1 SD unit increase in alcoholic drinks per week. PLT, platelet count. WBC, white blood cell count. Bars indicate 95% confidence intervals. *p<0.05.

Other genetically determined smoking traits, including smoking cessation (SmkCess), smoking heaviness (CigPerDay), and lifetime smoking score (LfSmk) did not significantly impact erythroid traits (**Supplementary Fig. 4**). SmkCess and CigPerDay were likely underpowered to detect any significant associations, although SmkCess (phenotypically opposed to SmkInit) trended toward slightly positive effects (**Supplementary Fig. 4**). However, LfSmk is a well powered phenotype (Wootton et al., 2019; Thom et al., 2020). Thus, it is possible that the slightly positive effects of LfSmk on erythroid traits reflect impacts from smoking cessation and/or heaviness that negate otherwise negative effects of smoking initiation genetics. The effects of LfSmk also agree with clinical observational data linking tobacco use with higher hemoglobin (Nordenberg et al., 1990; Malenica et al., 2017), which would inherently reflect smoking duration and heaviness that are sometimes unmeasured among clinical cohorts.

### Body mass index does not mediate the effects of SmkInit on erythroid traits

The negative effects of SmkInit on erythroid traits contrasted clinical observations, most of which would have anticipated positive effects of SmkInit on RBC, HGB, and HCT (e.g., (Nordenberg et al., 1990; Malenica et al., 2017)) We thus endeavored to identify possible confounding traits. For example, we previously identified body mass index (BMI) as a strong confounder for SmkInit (Thom et al., 2020) and a negative regulator of quantitative blood traits (Thom et al., 2022). However, our MVMR experiments argued against BMI as a confounder for the effects of SmkInit on erythroid traits, with consistent effect sizes and significance estimates for SmkInit on erythroid traits after adjusting for the effects of BMI, with some loss of power in the instrumental variable (**Supplementary Fig. 5**).

### Genetically determined alcohol consumption mediates the effects of SmkInit on erythroid traits

We next considered genetically influenced alcohol use as a potential mediating factor for the effects of SmkInit on erythroid traits, given clinical links between increased alcohol and tobacco use (Shiffman, 1996), as well as strong genetic associations between SmkInit and alcoholic drinks consumed per week (DrnkWk, r_g_=0.34, p=6.7×10^−63^, (Liu et al., 2019)).

Like SmkInit, an increase in genetically determined DrnkWk strongly negatively impacted HGB, HCT, and RBC across MR methods (**Fig. 1B, Supplementary Fig. 2** and **Supplementary Fig. 6**), without evidence of horizontal pleiotropy (all MR Egger intercept p>0.05). This was consistent with clinical observations related to the hematologic effects of alcoholism (Ballard, 1997).

We then confirmed using MR that an increased SmkInit risk portended an increase in DrnkWk across MR methods (**Fig. 2A**). Reverse causality and MR Steiger tests supported a unidirectional effect of increased SmkInit on increased DrnkWk (MR Steiger p-value 1.7×10^−67^, sensitivity ratio 3.1, **Fig. 2A**).

**Figure 2.**
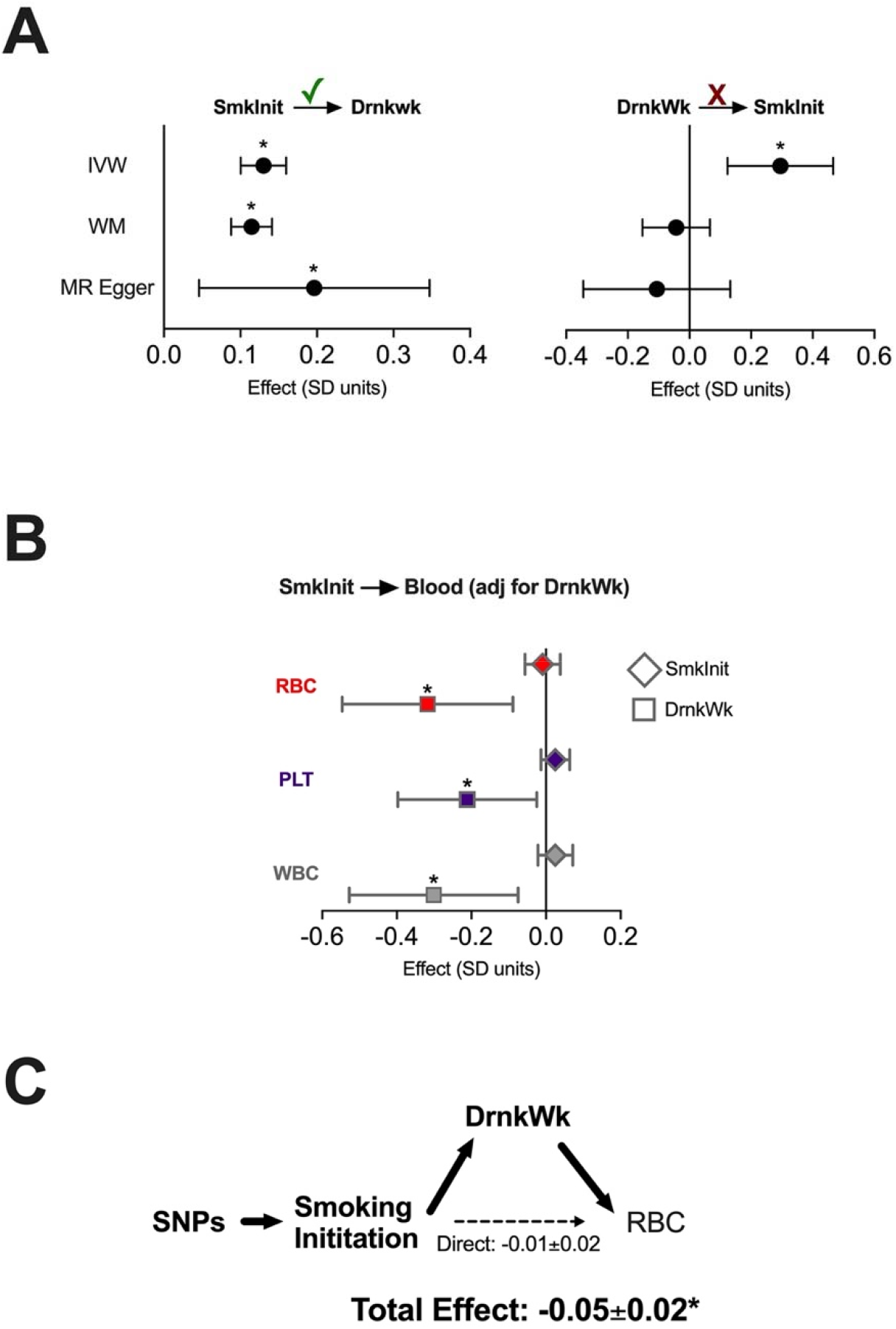
Genetically predicted alcohol use mediates the effects of smoking behavior on erythroid traits. **(A)** By MR, increased risk of SmkInit increases Drinks per week (Drinks) by inverse variance weighted (IVW), weighted media (WM), and MR Egger methods. However, increased genetically determined Drinks per week does not consistently increase SmkInit risk across MR methods. Check mark reflects MR Steiger ‘correct causal estimate’ relationship. **(B)** By MVMR, using an instrumental variable for SmkInit adjusted for Drinks per week, the effects of SmkInit on quantitative blood traits were nullified. Effects of Drinks per week on blood traits remain significantly negative. **(C)** Mediation analysis showed that SmkInit made a statistically insignificant effect on RBC, whereas the total effect (including indirect effects through Drinks per week) significantly negatively impacts RBC. Bars indicate 95% confidence intervals. *p<0.05.

In MVMR experiments, adjustment for DrnkWk abrogated the effects of SmkInit on erythroid traits (**Fig. 2B** and **Supplementary Fig. 7**). Formal mediation experiments confirmed that the effect of SmkInit on RBC was largely mediated indirectly through DrnkWk (**Fig. 2C**). The effects of LfSmk on erythroid traits were more positive after adjustment for DrnkWk, although did not reach statistical significance (**Supplementary Fig. 8**). However, multivariable results with LfSmk did reveal increased WBC, consistent with prior clinical and genetic studies (Sunyer et al., 1996; Pedersen et al., 2019). Thus, a genetic predisposition for increased alcohol consumption was an important confounding factor impacting the effects of tobacco use on blood traits.

## Discussion

Systemic factors and disease states have been clinically observed to alter blood cell formation and function (Heyde et al., 2021; Rohde et al., 2021). With few exceptions, systemic and environmental impacts on blood traits are not well characterized at a genetic level. The MR framework is a good way to assess genetic trait interactions, given its independence from confounding effects that can bias observational studies (Burgess et al., 2020). Here, we used MR to reveal causal effects of genetically determined tobacco and alcohol use on blood traits, providing evidence for novel trait associations.

Two aspects of these results were surprising. First, the effects of smoking were confined to erythroid traits. This contrasted the effects of obesity and adipose distribution, which impacted blood traits across lineages (Thom et al., 2022). While we previously inferred those findings to relate to hematopoietic stem and progenitor cell biology, the effects of tobacco and alcohol use seem mostly related to terminal erythroid maturation and/or erythrocyte homeostasis. This may have important implications for related genetic mechanisms that underlie these trait associations, which require further exploration to delineate. However, we anticipate that erythroid-specific mechanisms may target erythrocyte stability, splenic clearance, or renal function.

Second, the negative effects of SmkInit on erythroid traits countered prevailing clinical intuition and prior observations linking tobacco use with higher erythroid indices (Nordenberg et al., 1990; Malenica et al., 2017), although we note that our findings agree with prior studies linking smoking habits with anemia (Leifert, 2008). Our MVMR experiments revealed that these unexpected smoking trait relationships resulted from the co-inheritance of tobacco and alcohol use predispositions. However, our findings may reflect different biological mechanisms or context than most clinical studies, which have generally profiled long-term smokers. The increased genetic predisposition to initiate smoking, as defined by GWAS (Liu et al., 2019), does not necessarily equate to long term tobacco use nor all anticipated adverse health effects.

Consistent with this notion, our MR experiments using an instrumental variable for LfSmk showed no negative effects on erythroid traits, and trended toward a positive effect after adjustment for related factors. While these findings may have been complicated by underpowered SmkCess and CigPerDay effects, these findings may be more comparable to clinically observed association between tobacco use and higher erythroid indices.

Systemic and environmental factors can influence blood cell formation and blood cells impact myriad disease states. Understanding direct causal relationships between these factors is important. Given certain assumptions are met MR can determine genetic trait interactions free of the bias and confounders inherent to observational studies (Burgess et al., 2020). While future work is needed to further clarify exact loci and genes that link tobacco and alcohol use with blood trait variation, our results define novel causal effects of these behaviors on genetically determined erythroid traits.

## Methods

### GWAS collection

This study utilized publicly available GWAS data for tobacco and alcohol use (Liu et al., 2019), lifetime smoking score (n=462690) (Wootton et al., 2019), body mass index (N=484680) (Pulit et al., 2019), and blood traits (N=563085) (Vuckovic et al., 2020). All data were derived from individuals of European descent and used genome build hg19. Smoking initiation (SmkInit, N=1,232,091) was a binary phenotype derived from surveys querying whether an individual had ever smoked regularly for >1 month or having had smoking more than 100 cigarettes total in one’s life. Data for smoking cessation (SmkCess, N=547219), cigarettes smoked per day (CigPerDay, N=337334), and alcoholic drinks consumed per week (DrnkWk, N=941280), were also collected by report (Liu et al., 2019). SmkCess was defined in this publication as having consistently used tobacco previously, but having subsequently quit. Lifetime smoking scores (LfSmk), calculated from smoking initiation, duration, and heaviness, were based on UK Biobank data (Wootton et al., 2019).

### Genetic variant selection and instrumental variable creation

Instrumental variables were created by filtering summary GWAS statistics for SNPs common to exposure, outcoming, and/or mediating factor data sets. We used R v4.2.2 (R Development Core Team) and the package TwoSampleMR package (v0.5.6, (Hemani et al., 2018)) to clump SNPs meeting genome-wide significance for each exposure phenotype, selecting single SNPs in linkage disequilibrium (EUR r^2^<0.01) in 250kb genomic regions. Instrumental variable strength was estimated using Cragg-Donald F-statistics, instruments calculated to have an F-statistic>10 deemed to have limited weak instrument bias (Burgess and Thompson, 2011).

### Mendelian randomization and mediation analyses

Certain assumptions must be true to permit valid conclusions from MR studies, including that independent genetic instruments (SNPs) must be associated with the exposure trait. Weak instruments, horizontal pleiotropy, heterogeneity, and phenotype error measurements can limit applicability or inferences (Burgess et al., 2020). This study attempts to follow all best practices and reporting related to our MR experiments.

We used the TwoSampleMR package (v0.5.6, (Hemani et al., 2018)) to conduct univariable MR analyses. Univariable experiments focused on results from inverse variance weighted (IVW), weighted median, and MR Egger regression methods. We looked for evidence of horizontal pleiotropy using MR Egger regression intercepts; significant deviation from zero can imply directional bias (Bowden et al., 2015).

For MVMR experiments, we used the MVMR package (Sanderson et al., 2019). We report IVW causal effect estimates. From our MR Steiger analyses, we reported sensitivity, statistical significance estimates, and inference for correct causal direction (Hemani et al., 2017). In mediation experiments, we report total and direct effects estimates for each exposure and mediating trait on given outcomes (Burgess et al., 2017)

### Statistical analysis and data presentation

Presented effects include effect estimates from IVW, WM, and MR Egger regressions. Given heterogeneity in some instrumental variables, we utilized random effects models. Statistical significance was defined as p<0.05 for all experiments. Figures were prepared using GraphPad Prism 8.

## Data and code availability

Coding scripts and data sets, including full instrumental variables used in our study, can be found on Github (https://github.com/thomchr/SmkBloodMR and are also available upon request.

## Data Availability

Coding scripts and data sets, including full instrumental variables used in our study, can be found on Github (https://github.com/thomchr/SmkBloodMR) and are also available upon request.

## Acknowledgements

This work was supported by the National Institutes of Health (K99HL156052 to CST), University of Pennsylvania Undergraduate Translational Research Immersion Program (SS), and University of Pennsylvania Undergraduate Research Mentoring Program (VT).

## Supplementary Figures

**Supplementary Figure 1.**
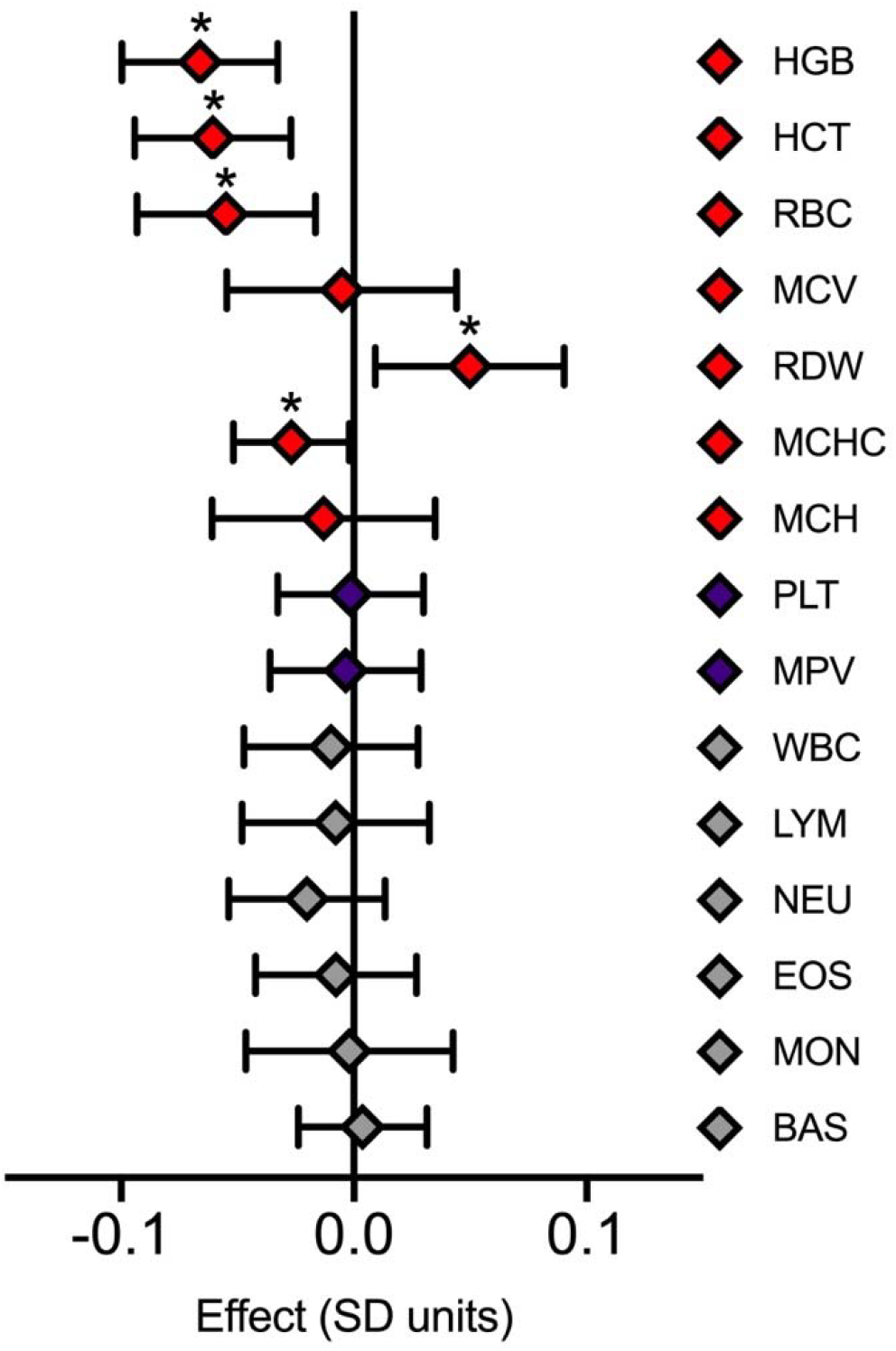
Univariable MR effect estimates for genetically influenced SmkInit on the indicated blood traits. Effects of a 2-fold increase in SmkInit risk on the indicated blood traits by univariable MR. Bars indicate 95% confidence intervals. *p<0.05.

**Supplementary Figure 2.**
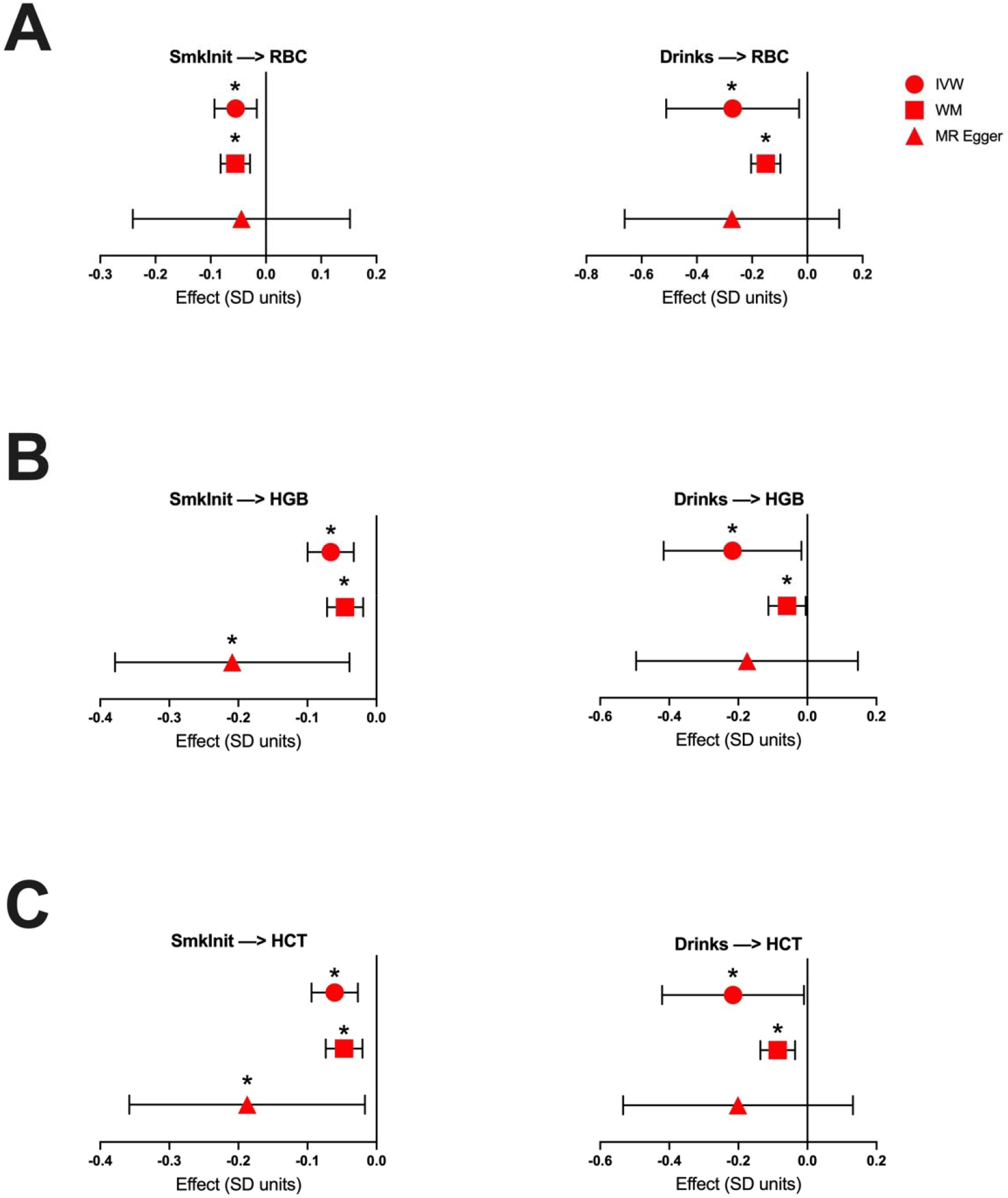
Sensitivity analyses for trait association in univariable MR experiments. (**A-C**) Effects of a 2-fold increase in SmkInit risk or a 1 SD unit increase in alcoholic drinks per week on (**A**) RBC, (**B**) HGB, or (**C**) HCT by weighted median (WM) and MR Egger regression analyses. IVW estimates also found in Figure 1 are presented here for comparison. Bars indicate 95% confidence intervals. *<0.05.

**Supplementary Figure 3.**
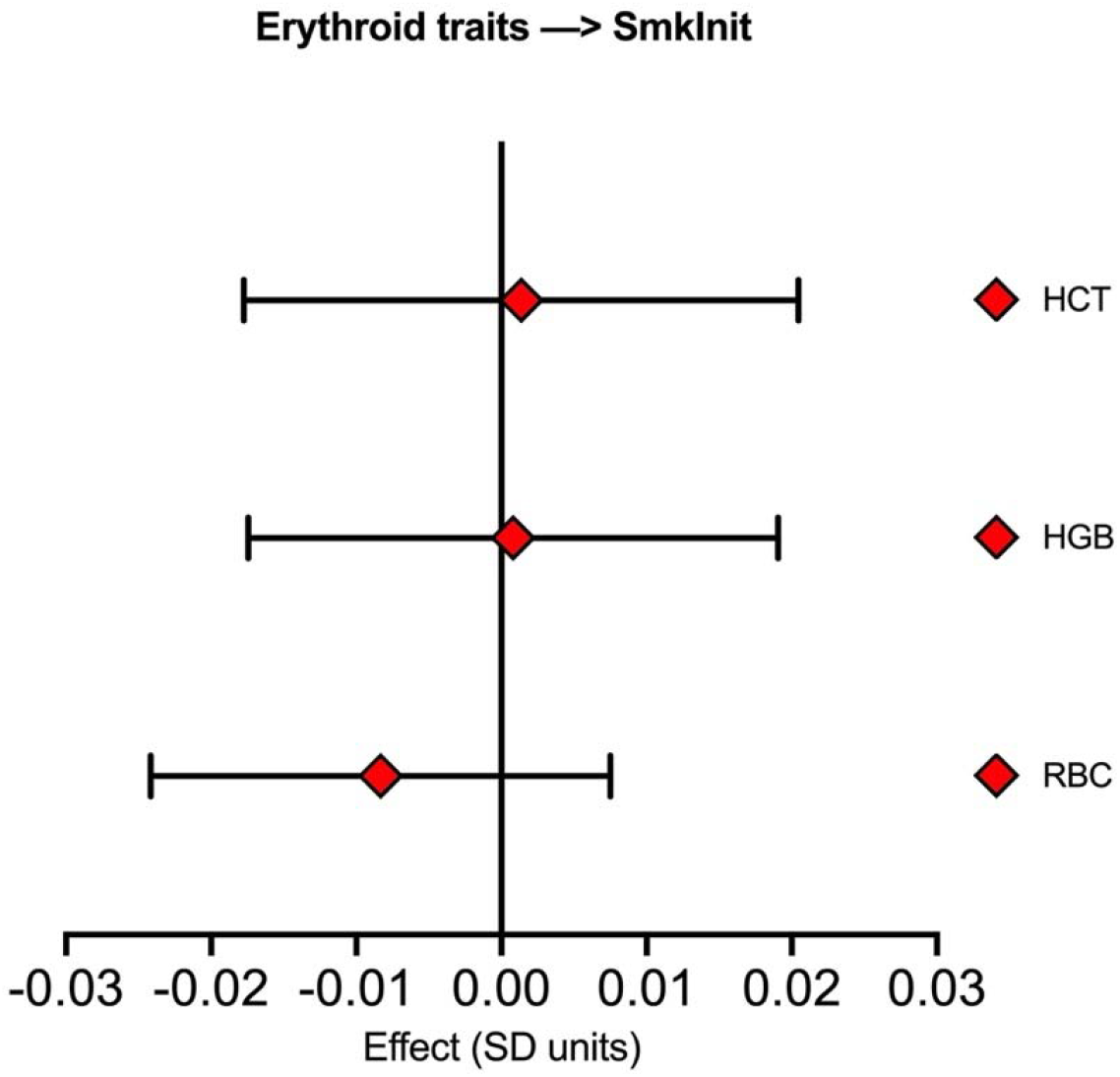
Univariable MR effect estimates for a 1 SD unit increase in the indicated erythroid traits on SmkInit. None of the estimates reached statistical significance. Bars indicate 95% confidence intervals.

**Supplementary Figure 4.**
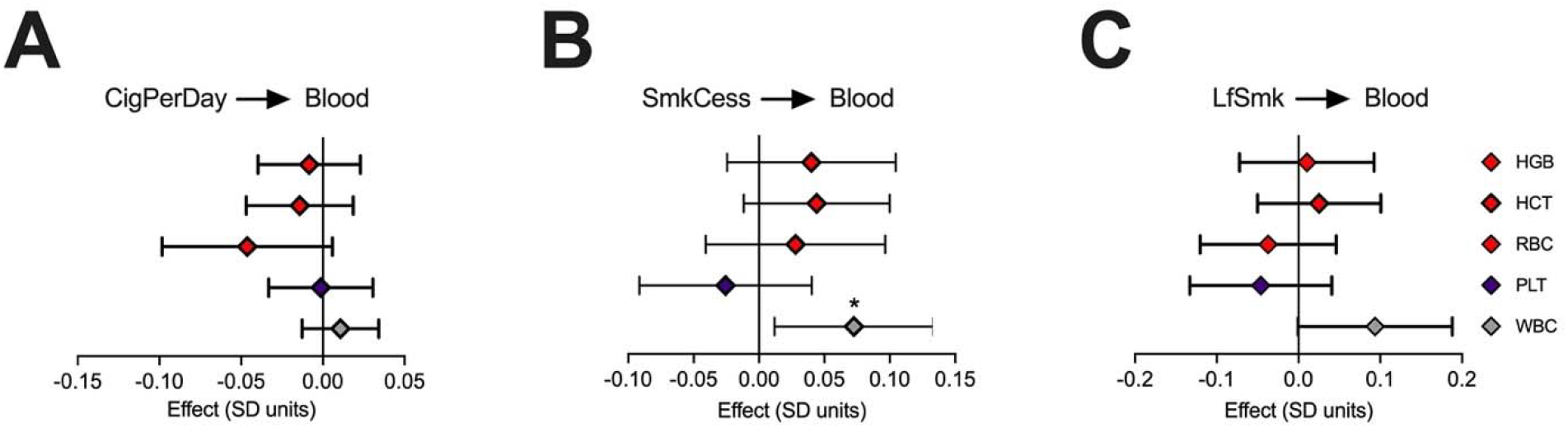
Univariable MR effect estimates for a 1 SD unit increase in smoking traits on the indicated blood traits. Bars indicate 95% confidence intervals. *p<0.05.

**Supplementary Figure 5.**
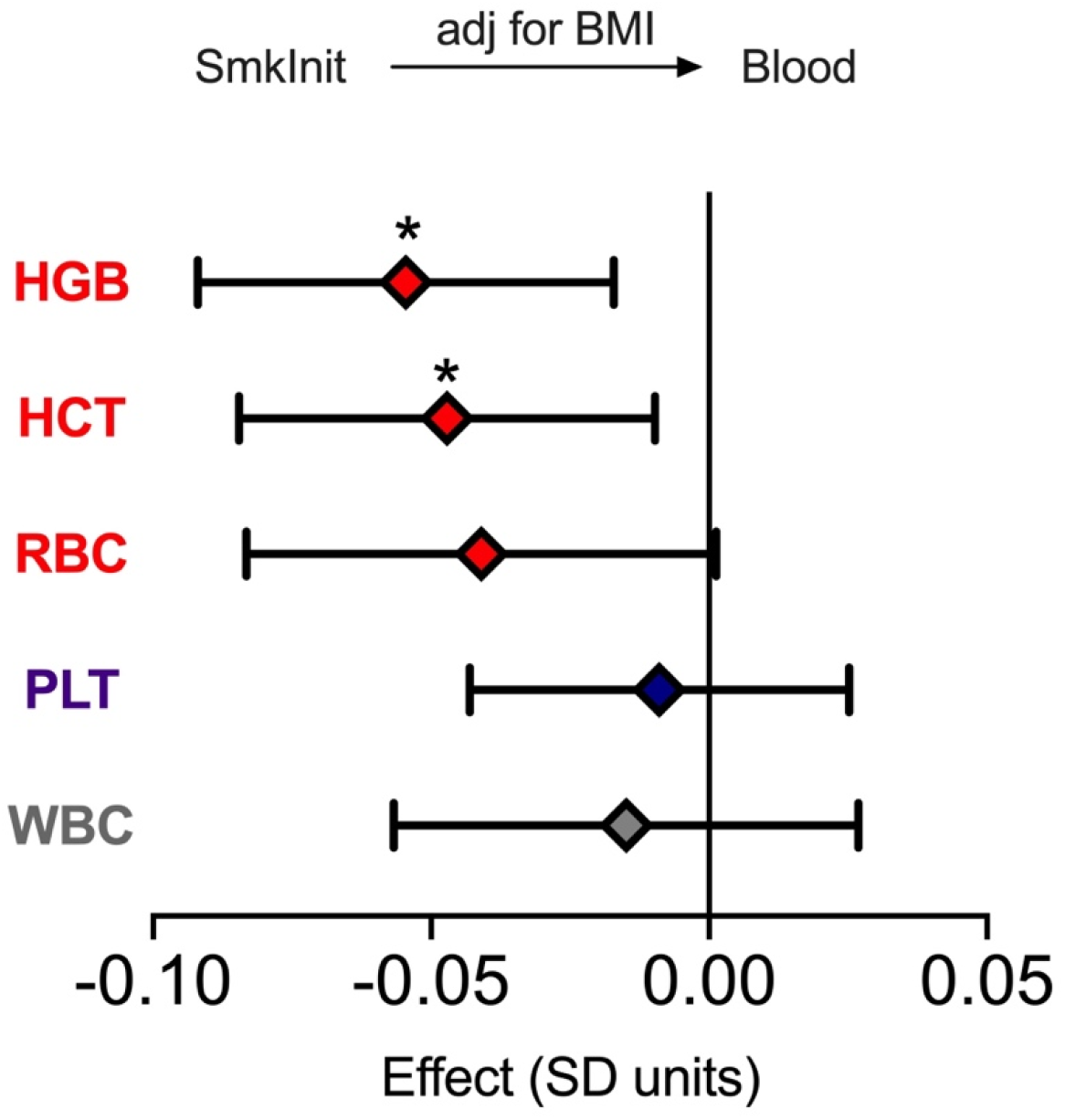
MVMR experiments analyzing the effect of SmkInit on the indicated blood traits after adjusting for body mass index (BMI). Bars indicate 95% confidence intervals. *p<0.05.

**Supplementary Figure 6.**
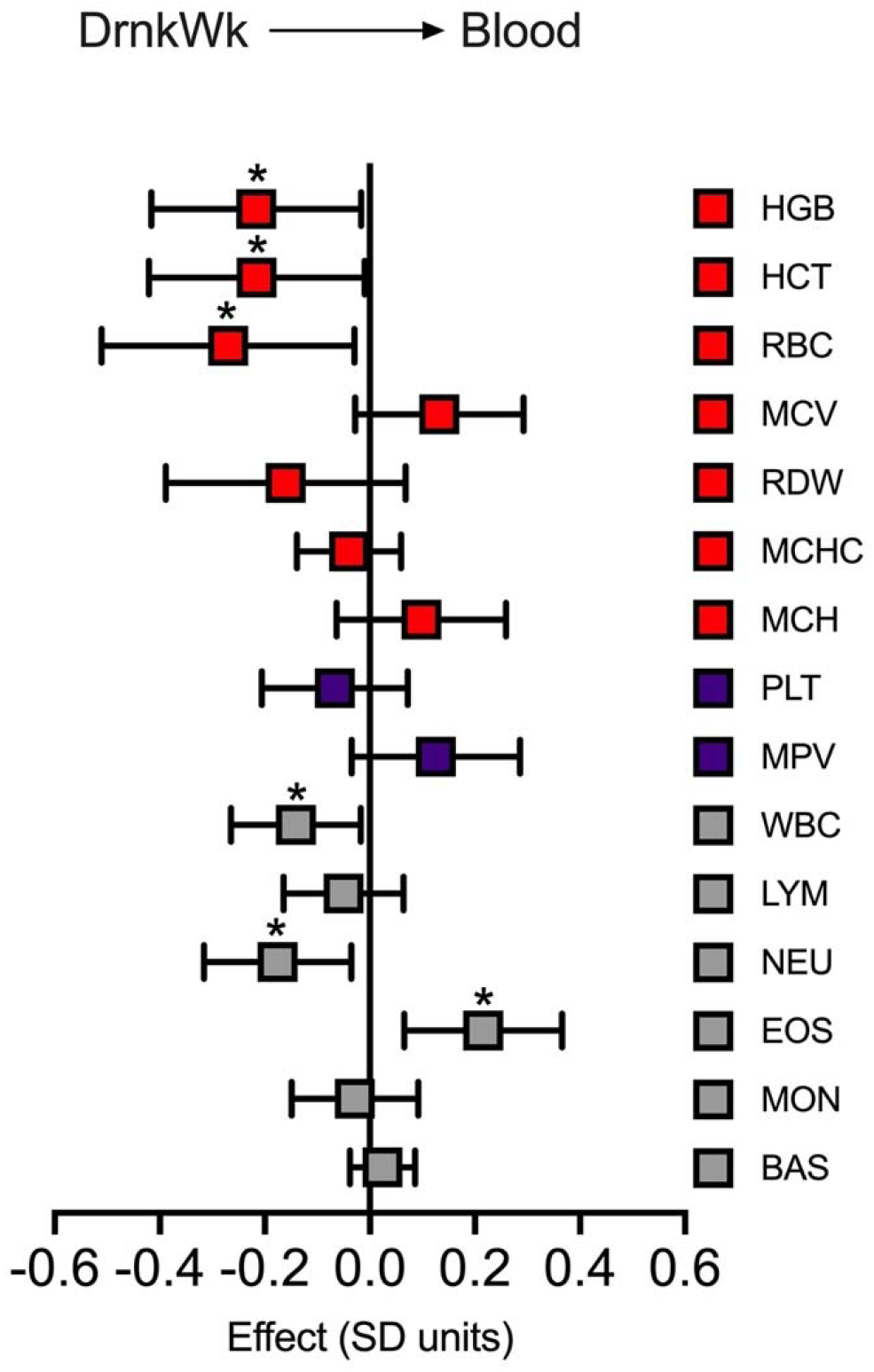
Univariable MR effect estimates for a 1 SD unit increase in DrnkWk on the indicated blood traits. Bars indicate 95% confidence intervals. *p<0.05.

**Supplementary Figure 7.**
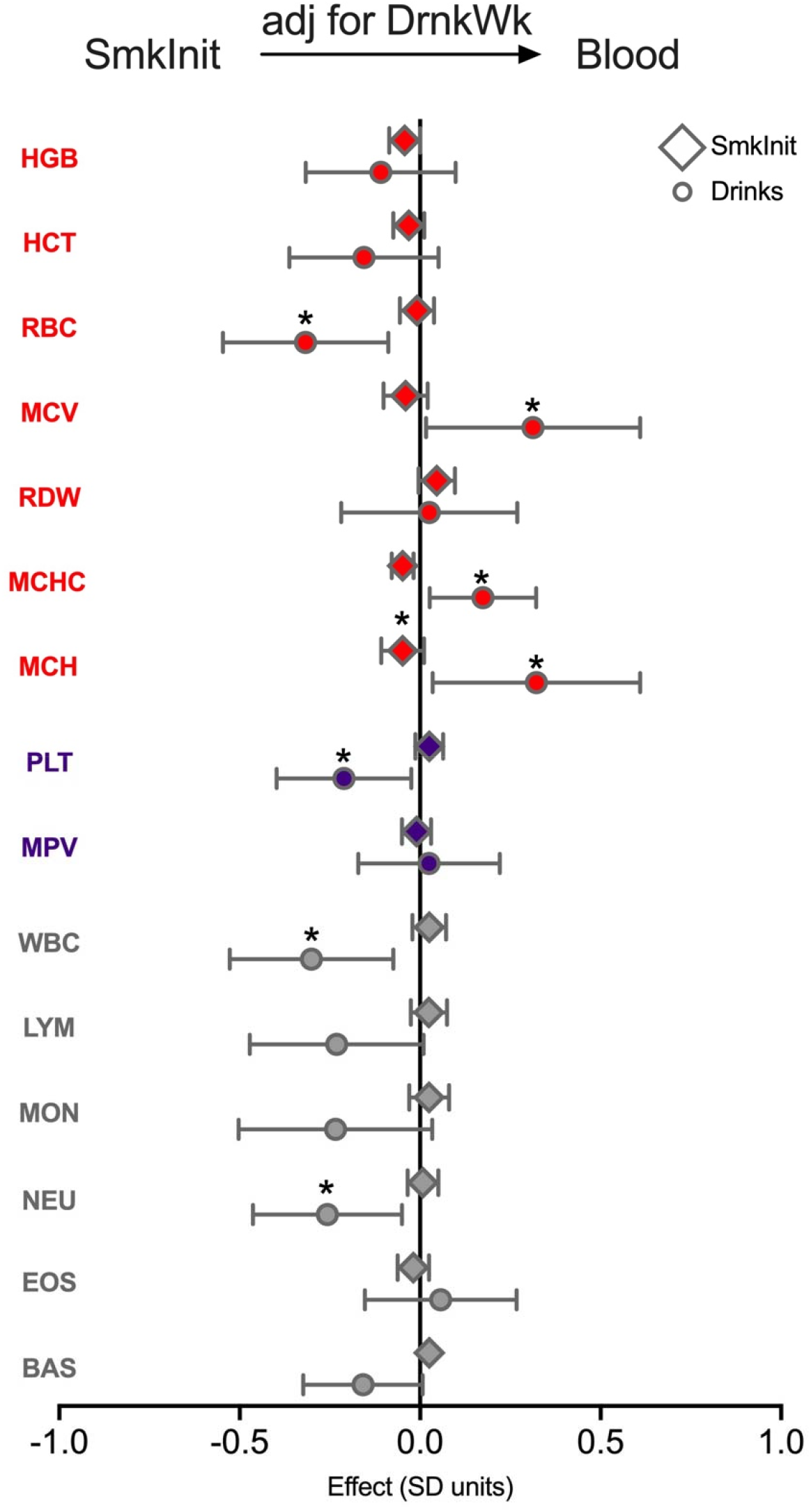
MVMR effect estimates for SmkInit or DrnkWk on the indicated blood traits. All experiments used an instrumental variable for SmkInit adjusted for DrnkWk. After adjustment, SmkInit did not have significant effects on any blood trait whereas DrnkWk did retain some significant effects. Bars indicate 95% confidence intervals. *p<0.05.

**Supplementary Figure 8.**
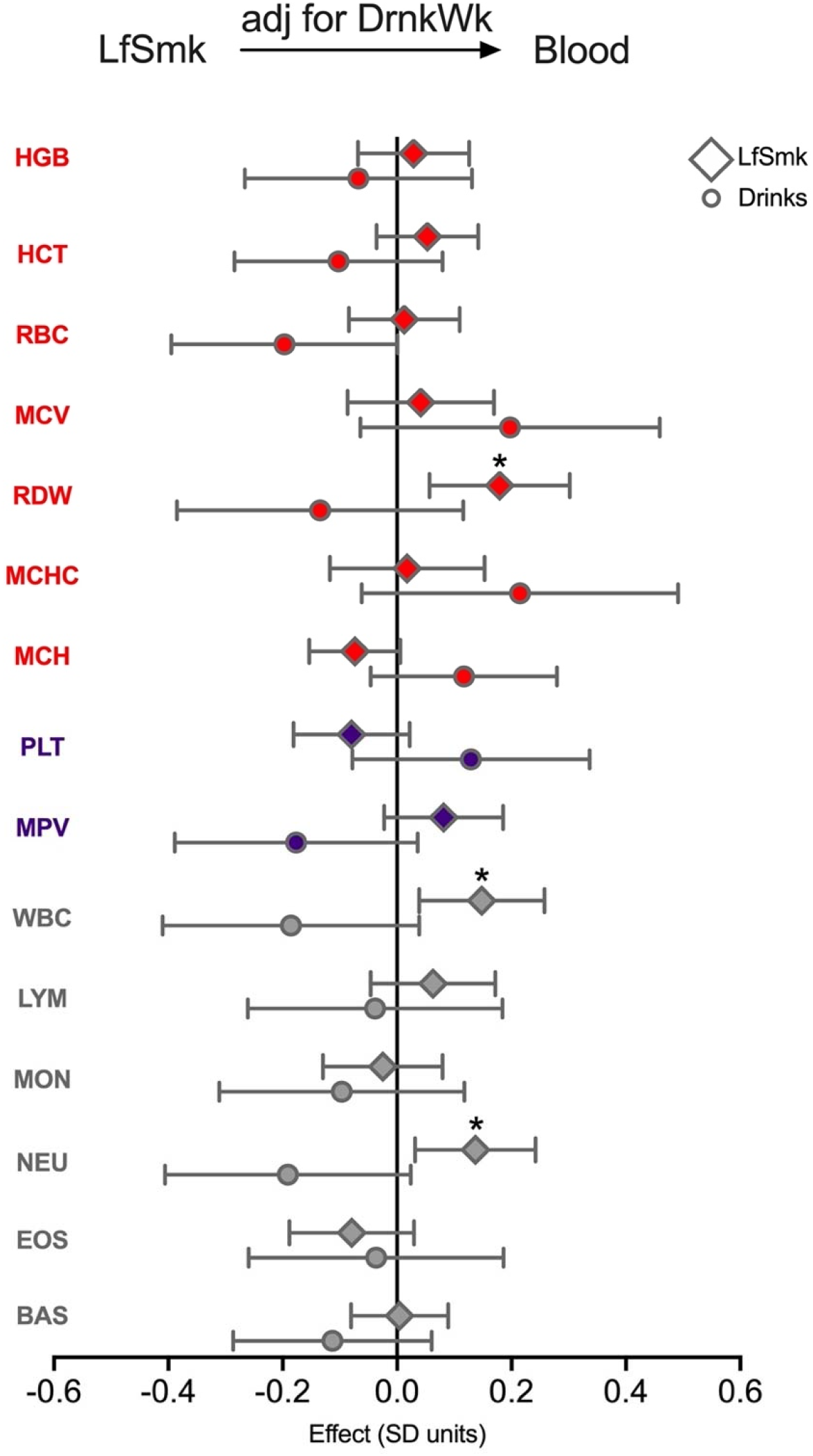
MVMR effect estimates for LfSmk or DrnkWk on the indicated blood traits. All experiments used an instrumental variable for LfSmk adjusted for DrnkWk. After adjustment, LfSmk did not have significant effects on any blood trait whereas DrnkWk did retain some significant effects. Bars indicate 95% confidence intervals. *p<0.05.

## Supplementary Tables

**Supplementary Table 1**. Explanations of genetically influenced traits analyzed in our study.

**Supplementary Table 2**. Craig-Donald F statistics for instrumental variables used in this study.

## Notes

### Competing Interest Statement

The authors have declared no competing interest.

### Author Declarations

This study used only publicly available GWAS summary statistics.

### Summary of Updates

This manuscript has been revised to include new information, structure, title and collaborators.

